# Portable Six-Channel Laser Speckle System for Simultaneous Cerebral Blood Flow and Volume Measurement with Potential Application for Characterization of Brain Injury

**DOI:** 10.1101/2024.10.30.24316429

**Authors:** Simon Mahler, Yu Xi Huang, Max Ismagilov, David Álvarez-Chou, Aidin Abedi, J. Michael Tyszka, Yu Tung Lo, Jonathan Russin, Richard L. Pantera, Charles Liu, Changhuei Yang

## Abstract

In regional cerebrovascular monitoring, cerebral blood flow (CBF) and cerebral blood volume (CBV) are key metrics. Simultaneous, non-invasive measurement of CBF and CBV at different brain locations would advance cerebrovascular monitoring and pave the way for brain injury detection, as current brain injury diagnostic methods are often constrained by high costs, limited sensitivity, and reliance on subjective symptom reporting. This study’s aim is to develop a multi-channel non-invasive optical system for measuring CBF and CBV at different regions of the brain simultaneously with a cost-effective, reliable, and scalable system capable of detecting potential differences in CBF and CBV across different regions of the brain. The system is based on speckle contrast optical spectroscopy (SCOS) and consists of laser diodes and board cameras which have been both tested and investigated for safe use on the human head. Results on a cohort of five healthy subjects indicated that the dynamics of both CBF and CBV were synchronized and exhibited similar cardiac period waveforms across all six channels. As a preliminary investigation, we also explored the potential use of our six-channel system for detecting the physiological sequela of brain injury, involving a subject with significant structural brain damage compared to another with lesser structural brain damage. The six-point CBF and CBV measurements were compared to MRI scans, revealing that regions with altered blood dynamics closely correlated with the injury sites identified by MRI.

## 1 Introduction

Brain injury can occur from traumatic and non-traumatic mechanisms. Traumatic brain injury (TBI) is one of the leading causes of death and disability among young people worldwide^1–3^. A TBI occurs when the brain experiences excessive non-physiological mechanical forces which can lead to hemorrhage, contusion, inflammation, cell death, edema, and/or ischemia. After recovery from TBI, patients can exhibit persistent evidence of structural brain damage as seen on MRI, as well as physiological disruptions such as cerebrovascular dysregulation that lead to subtle functional deficits that may worsen with time if left untreated^4,5^. While structural sequela of TBI can be readily characterized by MRI, objective measures of cerebrovascular reactivity and cerebral blood flow can be very helpful in fully characterizing the effect of TBI^3^, including mild cases not associated with obvious structural damage to the brain. The incidence of mild TBI with minimal structural brain damage is particularly high - there are nearly three million mild TBI occurrences in the US each year with the majority occurring in adolescents and young adults. Mild TBI is also one of the leading causes of injuries in the U.S. Army, with blast-related TBI often described as the signature injury during deployment^6–8^. Despite the heavy injury toll and almost two decades of research, the diagnosis, treatment, and recovery from TBI remains poorly understood. Various methods have been studied for characterizing TBI, including MRI-based neuroimaging^9^, electrophysiology^3,10^, blood and saliva biomarkers^11–13^. Beyond TBI, non-traumatic brain injury (NTBI) can also lead to structural and physiological sequela including conditions such as stroke, hypoxia, infections, and toxic exposures. These injuries can result in damage to brain tissue, leading to long-term cognitive, motor, and sensory deficits depending on the affected brain regions. New methods that are cost-effective, comprehensive, and reliable enough for practical clinical use are necessary to better characterize brain injuries beyond structural damage, particularly the persistent disruption of normal cerebrovascular reactivity and cerebral blood flow^14^.

Recently, laser speckle contrast imaging was applied for monitoring cerebral blood flow (CBF) and cerebral blood volume (CBV) non-invasively on humans^15–20^. The technique, commonly called speckle contrast optical spectroscopy (SCOS)^16–21^ or speckle visibility spectroscopy (SVS)^15,21–25^, uses an infrared laser source to transmit light through the skull and brain in humans, Fig. 1(a). By transmitting infrared light through one location on the skull and collecting its transmission with a camera on another location, it is possible to determine brain blood volume by measuring the light attenuation rate^17,21^. Since the light used is also coherent (laser), it is possible to also determine brain blood flow rate by recording how fast the transmitted speckles fluctuate.

**Fig 1.**
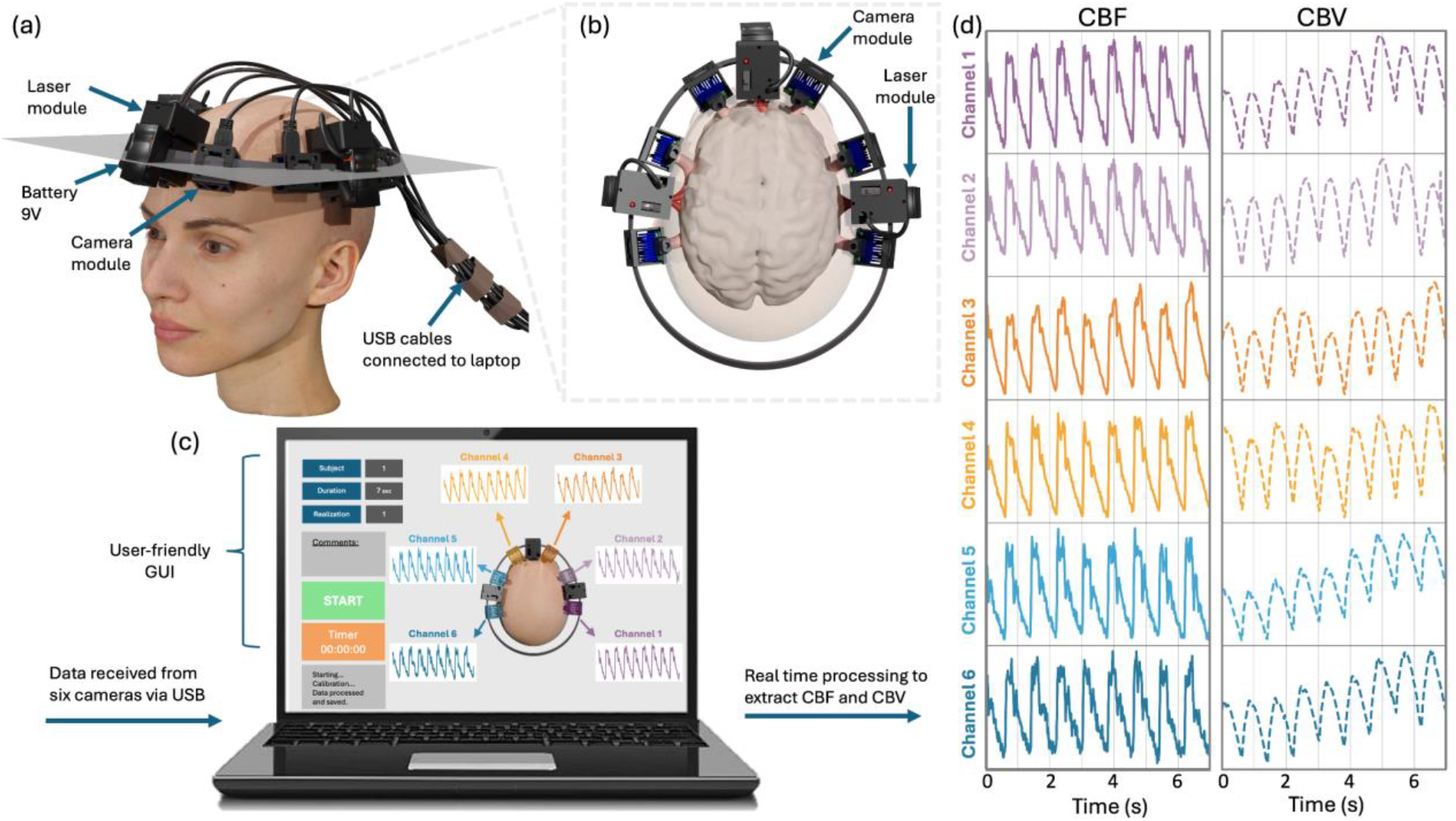
Experimental arrangement of the six-channel SCOS system. (a) 3D visualization of the six-channel system positioned on the head. (b) Schematic of the light penetrating the brain. (c) Graphical user interface for operating the system and for real time visualization of the cerebral blood flow (CBF) and cerebral blood volume (CBV). (d) Typical CBF and CBV blood dynamics measured at six different locations of the head from our compact SCOS system.

In this paper, we present a six-channel compact and portable SCOS system capable of simultaneously measuring CBF and CBV at six different brain locations (Fig. 1). The system includes three laser diode modules and six USB board camera sensor modules, which can be conveniently positioned around the subject’s head at distinct locations. Apart from the camera cables connection to a laptop, the entire system is integrated into a wearable headband and is powered by embedded and rechargeable 9V batteries. Data is transmitted via USB for real-time processing, enabling operation in diverse environments. The camera modules are designed to dissipate heat effectively, ensuring a comfortable contact temperature during extended recordings. Additionally, a graphical user interface (GUI) enables non-experts to operate the portable six-channel SCOS system with ease. We characterized and tested the system on a cohort of five healthy human subjects. The results showed synchronized and highly correlated dynamics of blood flow and blood volume across all six channels. Comfort evaluation tests confirmed the wearable system’s comfort during recording and established the system’s maximum operational duration.

As a preliminary investigation, we explored the potential use of our six-channel system for detecting the persistent effects of TBI using time-synchronized measurements of blood flow and blood volume at different brain locations, which offer a physiology-based approach to assess regional CBF and CBV differences, with suspected injured regions displaying different blood dynamics. We conducted a preliminary study involving two subjects who have a history of brain injury, one traumatic and the other non-traumatic. Each of the two subjects have undergone decompressive hemicraniectomy and cranioplasty, but with different degrees of persistent structural damage to the brain and compared their results with those of the five healthy subjects. The six-point CBF and CBV measurements were then compared to MRI scans, revealing that regions with altered blood dynamics closely correlate with the locations of structural damage identified by MRI.

## 2 Methods

### 2.1 Six-channel SCOS system

The arrangement of our compact six-channel SCOS system is shown in Fig. 1. The wearable headband and location of each channel on the head are shown in Fig. 1(a). The system contains three laser sources for illumination and six board-cameras for detection. The cameras are distributed on the head as follows: two on each side of the forehead, one on the front and one on the back of the left hemisphere, and one on the front and one on the back of the right hemisphere. Each camera is placed at a source-to-detector (S-D) distance from the laser source around 3.2 ± 0.2 cm, see Fig. 1(b). It was previously reported that a S-D distance between 3.0 to 3.5 cm corresponds to an optimal brain sensitivity over signal-to-noise ratio^15,21,26– 30^. We note that, even at such a large S-D distance, the collected signal would still be influenced by blood flow in the scalp. Each laser source is a single-mode continuous wave laser diode of 808 nm [Thorlabs M9-808-0150]. To ensure control over the illumination spot size and prevent undesirable laser light reflections or stray light, the laser diode is housed in a 3D-printed mount with a circular aperture of 5.5 mm diameter. A sliding block switch at the end of the laser mount can block laser light when the system is not in use. The camera is also housed in a 3D-printed mount. All the mounts were printed using an Anycubic 3D printer in black resin which absorbs light and minimizes back reflection and stray light disturbances. The laser diode is positioned 6 mm away from the skin of participants such that the illumination spot diameter was 5.5 mm^15,16^. The total illumination power is limited to 67 mW to ensure that the laser light intensity level of the area of illumination is well within the American National Standards Institute (ANSI) laser safety standards for maximum permissible exposure (3.28 mW/mm^2^) for skin exposure to a 808 nm laser beam^31^. Each laser is operated by a custom-made printed circuit board combined with a laser diode driver [Thorlabs LD1100], both encased atop the laser diode, see Fig. 1(a). Three 9V batteries (located on the top area of the head) provide electrical energy for switching the three laser sources with battery life lasting greater than 3 hours of usage (Figs. 2(a) and 2(b)).

**Fig 2.**
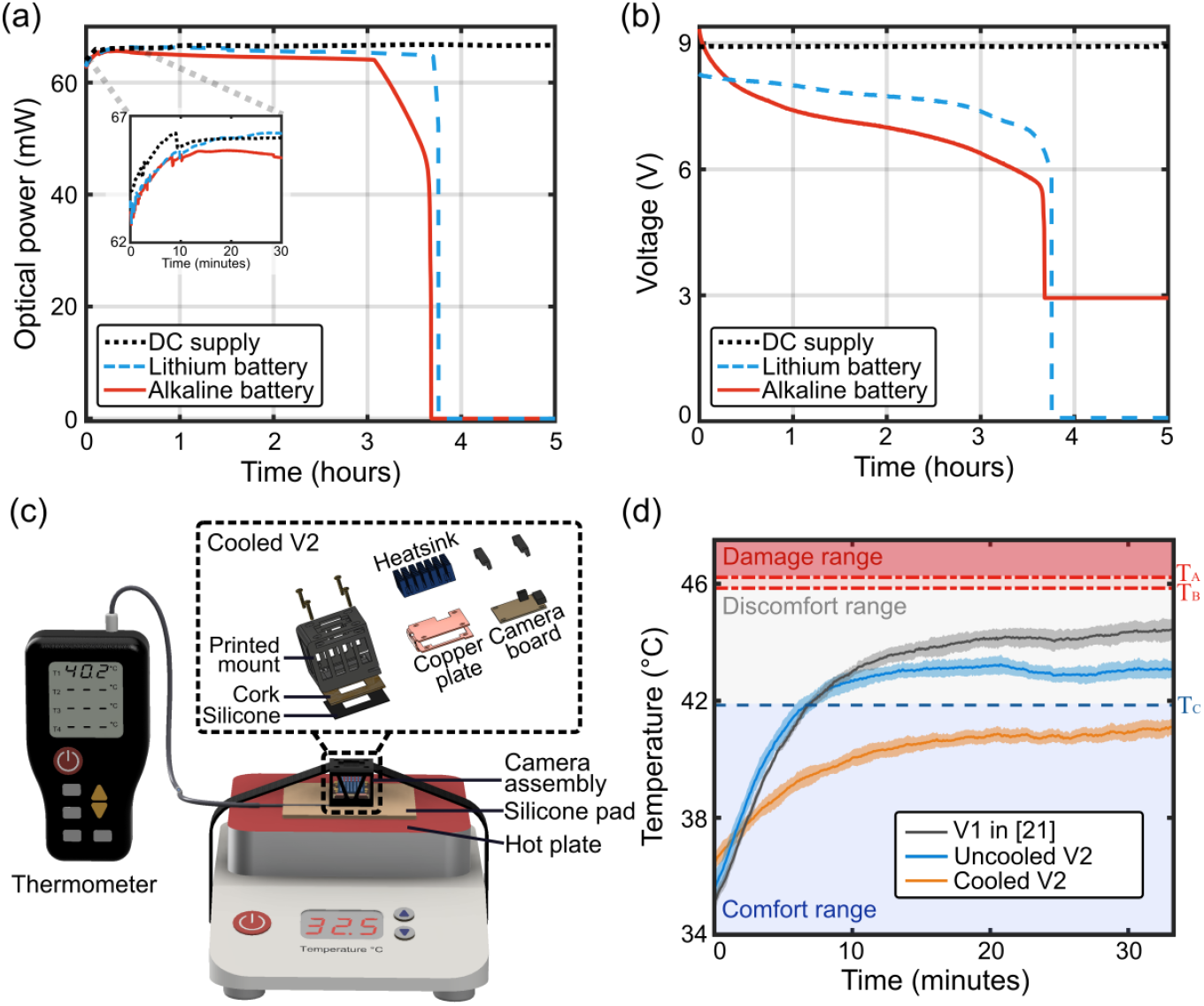
Optical lasing power and camera heating temperature characterization of the system. (a) Optical power and (b) power supply voltage as functions of time. (c)

The camera used is a rolling shutter camera [Basler daA3840-45um (Sony IMX334 sensor)] operating at a framerate of 40 frame-per-second, with a pixel pitch of 2×2 μm, a pixel resolution of 3840 × 2160 pixels, a pixel depth of 8 bits, and operated at an exposure time *T* = 6 ms. A heat management system ensures that the contact temperature of the camera mount remains below the safety threshold. The passive heat management system comprises of the following materials: a 0.040” laser-cut copper sheet, a 1 mm neoprene sheet [McMaster 93375K608] and an anodized aluminum heat sink of 30×20×10 mm placed on the back of the copper sheet, a 1 mm thickness cork sheet, and a 1 mm thickness black silicone sheet. The design and dimensions of the camera module system are illustrated in Fig. 2(c).

### 2.2 CBF and CBV measurements

The estimated number of speckles per pixel collected by the camera is about 4 speckles per pixel, corresponding to a one-dimensional speckle-to-pixel length ratio of s/p = 0.5^24,32,33^. The speckle contrast K is calculated from the recorded speckle images as:

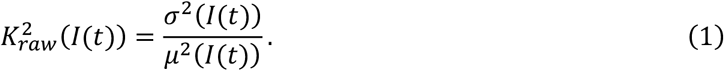

Where *σ*^2^(*I*(*t*)) is the variance of the normalized image *I* at time *t* is and *μ*(*I*(*t*)) its mean. The various source of noises accounted as^16,18,34,35^:

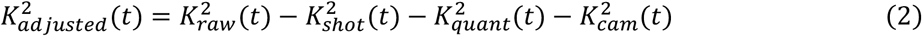

with 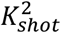 accounting for variance contributions from the shot noise, 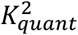 for the variance inherited from quantization, and 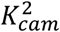 for the variance contributions of the camera’s readout noise and dark noise. See Ref^16^ for more details about the speckle contrast calculations and calibration processes. The cerebral blood flow index (CBFI) is related to 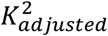 as^16^:

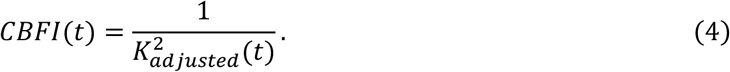

The blood volume is extracted from the camera images by calculating the cerebral blood volume index (CBVI) from the recorded images as^17,21^:

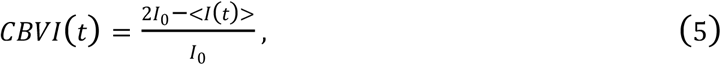

where *I*_0_ is the intensity at baseline. In the results of this paper, we utilize CBFI and CBVI metrics to provide normalized blood dynamics information for enhanced comparability across measurements.

The graphical user interface, illustrated in Fig. 1(c), controls the system, and displays the CBF and CBV results at the end of each recording. The Python-based GUI prompts users to input details such as subject information, trial number, and the duration of the recording. The GUI caps the recording duration to 180 seconds. Users can also add comments regarding the qualities of the recording, potential movements, and subject information. Furthermore, the GUI ensures safety by requiring users to confirm adherence to all laser safety protocols before initiating measurements. Upon completion, the GUI immediately presents the results, including CBF and CBV for each channel, Fig. 1(c). The real-time processing of data negates the need to save raw images, thereby optimizing disk space usage. The total cost of the six-channel SCOS system hardware is about $3,000. The data collection and processing are performed by a Nvidia RTX 4070 laptop (∼$1,500).

Figure 1(d) shows typical CBF and CBV time traces measured over 5 seconds by the six-channels SCOS system on a human subject. As shown, all CBF and CBV time traces exhibit the same frequency of oscillation and are temporally synchronized. Note that the CBF time traces contain more high frequency information than the CBV time traces, including variations such as the peak systole or dicrotic notch of the cardiac pulse^21^. Additionally, the CBF time traces show greater temporal stability compared to CBV, as minor fluctuations in blood oxygen concentration and other instabilities do not significantly impact the normalized speckle contrast calculations (Eq. 1).

### 2.3 Battery-Powered Laser Source and Heat Management in Camera Modules

Next, we evaluated the stability and longevity of the laser source when powered by a DC power supply versus a 9V battery, Figs. 2(a) and 2(b) and examined the contact temperature of the camera module, Figs. 2(c) and 2(d). These characterizations are critical for using the system in a clinical setting, as patients in such settings may not be able to respond to pain or discomfort, or they may be unconscious. Therefore, it is essential to first ensure that powering the system with a low voltage battery guarantees electrical safety for the patients being scanned, and that the system’s temperature is carefully constrained to prevent any risk of heat-induced damage.

Figure 2(a) shows the laser output power, measured with an optical power meter positioned on the mounted laser diode, for three different power sources: a 9V DC bench power supply, a 9V alkaline battery, and a rechargeable 9V lithium-ion battery.

As shown, all three power sources provided the laser with an optical power of approximately 66 mW over time. The DC power supply maintained a constant output, while the alkaline and rechargeable lithium-ion batteries’ power decreased to zero after a few hours due to discharge. Overall, the lithium-ion battery outperformed the others, lasting the longest (approximately 3.7 hours), demonstrating the highest stability in optical power output, and being the most lightweight. Detailed performance comparisons of the batteries can be found in Appendix A. Additionally, the system optical power with the rechargeable lithium-ion battery sharply dropped to zero within 3 minutes, due to an internal electrical switch that automatically disables the battery when the voltage falls below a certain threshold. In our case, this feature is beneficial as it prevents noisy data collection during the battery’s discharge phase. With an average measurement time of 15 minutes per subject, the lithium battery can support up to 12 recordings or more on a single charge. This is practical, as the battery can be recharged between sessions.

Next, we tested the effectiveness of a passive heat management system for the camera modules, by measuring the contact temperature after placing the system on silicone skin pads [Stylia] positioned on a hot plate [VWR] set at 32.5 °C to simulate human skin contact (Fig. 2(c)). Our goal was to ensure compliance with thermal guidelines for skin contact (ASTM C1055^36^ and IEC60601^37^) and to provide user comfort during extended system operation. From the ASTM guidelines, we established three temperature thresholds for one hour of operation of the system: 1) a comfort temperature threshold T_A_ = 41.85 °C, 2) a reversible skin damage threshold temperature T_B_ = 45.85 °C, and 3) a severe skin damage threshold temperature T_C_ = 46.22 °C. Our aim was to not overcome the comfort temperature T_A_. We tested three camera modules. The first (V1) was the camera module [Basler daA1920-160um] used in Refs.^16,21^ running at 80 FPS with 2.3 million pixels, the second (uncooled V2) was a Basler daA3840-45um board camera running at 30 FPS and 8.3 million pixels. The third (cooled V2) was the camera module we used in the rest of this paper. It consists of a camera module with Basler daA3840-45um board camera running at 40 FPS and 8.3 million pixels encased in the heat management system shown in Fig. 2(c). For heat management, we attached an anodized aluminum heatsink to a C-shaped copper plate containing the camera board. On the front side, we incorporated a 1 mm layer of cork and a 1 mm layer of neoprene to further insulate heat. For experiments involving human subjects, we replaced the neoprene sheet with a silicone sheet to allow for easier and more thorough sanitization. The averaged temperature results for each module are presented in Fig. 2(d). As shown, all the cameras’ temperatures remained below both the reversible T_B_ and irreversible T_C_ damage thresholds, in accordance with ASTM guidelines. However, only the cooled V2 module stayed within the targeted comfort temperature range T_A_.

### 2.4 Human research study

Participants for this study were recruited from the greater Los Angeles areas, selected from adult volunteers aged 18 to 70 years. Before the experiments, each participant completed a health questionnaire, and their blood pressure was recorded. Informed consent was obtained from all participants beforehand. The human research protocol was approved by the Caltech Committee for the Protection of Human Subjects and the Institutional Review Board (IRB), Caltech IRB Protocol IR21-1074. To simplify the experimental setup and system implementation, measurements were taken on hairless areas or areas with little hair. The total illumination power adhered to the American National Standards Institute (ANSI) laser safety standards for maximum skin exposure to an 808 nm laser beam.

A total of seven subjects were enrolled in the study, including five “healthy” subjects with no history of brain injury. In addition, two subjects with a history of severe brain injury were also recruited. One “TBI” subject had a history of severe TBI, decompressive hemicraniectomy and subsequent cranioplasty skull implant (SI). The other “NTBI” subject suffered non-traumatic brain injury from large brain hemorrhage due to a ruptured arteriovenous malformation. The “NTBI” patient also underwent decompressive hemicraniectomy and subsequent cranioplasty. The “NTBI” patient had more evidence of structural brain damage and was still in post-discharge care due to behavioral issues at the time of the study. The “TBI” patient had relatively less structural brain damage and was living independently. Neither had focal neurological deficits.

## 3 Results

For blood flow measuring systems, characterizing the system’s contact temperature during operation is critical to ensuring participant safety. This is particularly important for systems in use in clinical settings, where patients may be unable to respond to pain or discomfort or may be unconscious^38^. While the laser source of our system adheres to ANSI standards, limiting the optical power level to prevent heat induction in participants, the detection module, working at a high speed, generally generates heat^39,40^. In systems where the modules are in direct contact with the participants’ skin, heat becomes a significant concern^38–40^, especially since these units typically operate at high speeds. To mitigate this issue, detecting devices can be operated for only a short period of time as short as a few minutes, or are switched on and off to allow for cooling between scans. Although these methods provide partial solutions, they are not practical for non-expert users and still pose safety risks. To address these challenges, we have designed a passive heat modulation system, as illustrated in Fig. 2, enabling our system to operate for more than half an hour while maintaining a comfortable temperature range. Passive cooling offers the advantage of regulating temperature without the need for complex or vibrating equipment with active external energy sources, making it more user-friendly and more practical for long-term use^40,41^.

In Fig. 3, we measured the contact temperature of our detecting module on the foreheads of five human subjects, with the camera running for 35 minutes with repeated 3-minute image acquisition sessions followed by 30-second breaks in between each session. This approach simulated an extreme usage scenario that may occur during hospital scanning. The room temperature was maintained at approximately 23°C to mimic typical hospital conditions, where temperatures are usually kept between 20°C and 24°C. The temperature was recorded using a commercial thermocouple [Risepro 4 channels], positioned between the camera module and the participants’ foreheads. Across recordings from all five subjects, the temperature stabilized after 20 minutes and never exceeded 41 °C, remaining within the comfort temperature range. These results are consistent with those shown in Fig. 2(d) from the silicone hotplate tests. Participants were also asked to provide subjective comfort ratings on a scale of ‘Comfortable’, ‘Mild’, ‘Tolerable’, ‘Painful’, or ‘Severe’ throughout the 35-minute experiment, Fig. 2(d). None of the subjects reported intolerable discomfort during the entire duration of the study. Only one participant indicated a comfort level worse than mild, which we attributed not to heat but to the pressure of the system due to its tightening. These experiments demonstrate that the multichannel compact SCOS system can operate for extended durations without the risk of heat-induced skin damage while maintaining an adequate level of user comfort.

**Fig 3.**
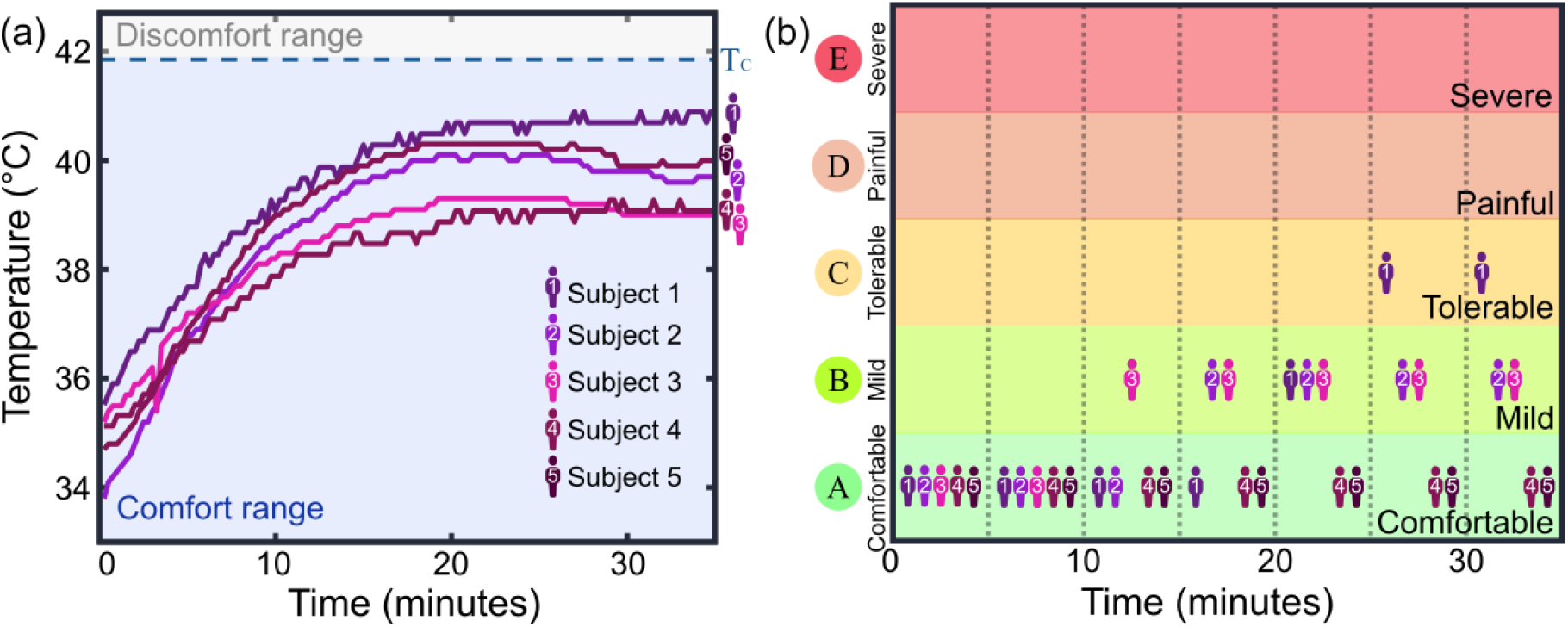
(a) Temperature measurements taken at the contact surface of the camera module onto the foreheads of five human subjects. The measured temperature saturated after twenty minutes of continuous operation, and always remained within the comfort range (i.e. below T_A_ = 41.85 °C). (b) Comfort ratings provided by participants during the 35 minutes of continuous recording. The room temperature was 23°C.

Using the six-channel portable system, we measured CBF and CBV on a cohort of seven subjects. The cohort consisted of five subjects with no history of brain injury (healthy), one subject with a cranioplasty skull implant (TBI) following a TBI but no major structural brain damage at the time of the study, and one subject with non-traumatic brain injury and undergoing cranioplasty skull implant at the time of study in post-discharge care (NTBI). Figure 4 shows the typical CBF measured on a healthy subject (Fig. 4(a)), the TBI subject (Fig. 4(b)), and the NTBI subject (Fig. 4(c)). As shown, the CBF data for the healthy subject displays synchronized and highly correlated blood flow dynamics across all six channels. A similar pattern is observed in the TBI subject. MRI scans confirmed that neither the healthy subjects nor the TBI subject had significant brain damage. The CBF data for the NTBI subject exhibited synchronized and correlated blood flow dynamics across all channels except the two channels positioned over the area of brain injury revealed by the MRI scan.

**Fig 4.**
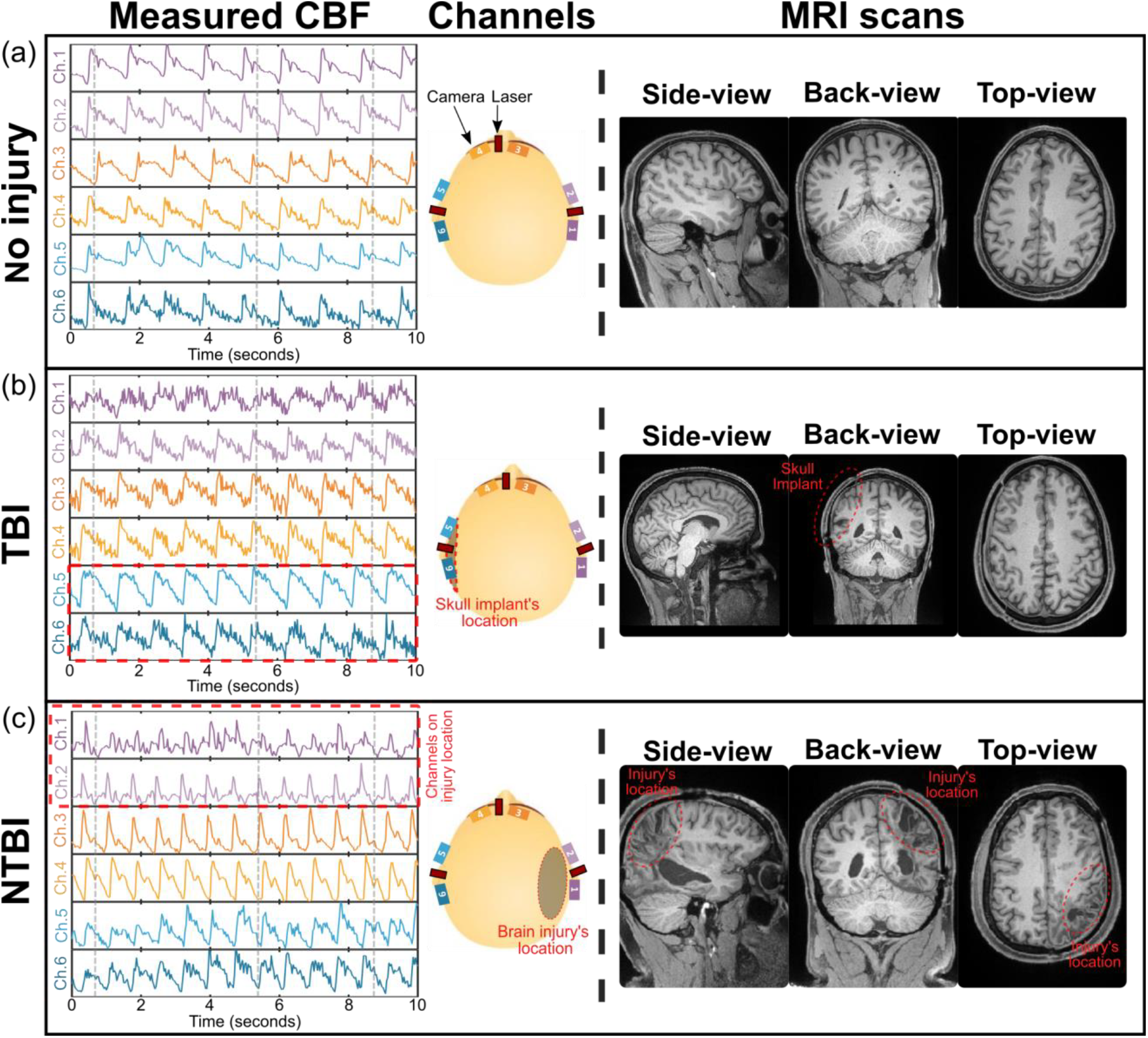
Measured CBF at six different locations and MRI scans of the heads of (a) healthy, (b) TBI, and (c) NTBI subjects. Note that the MRI images are not mirrored; the right side of the image corresponds to the subject’s right side.

To further characterize the differences across channels, we calculated the mean intensity of each of the six channels from the CBV recordings (Fig. 5(a)) and the two-channel correlation factor from each subject’s CBF recordings (Fig. 5(b)). The two-channel correlation factor was calculated using the Pearson correlation formula as:

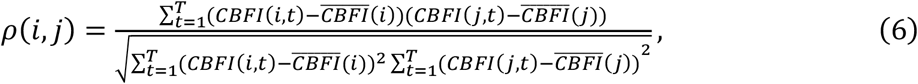

where *CBFI*(*i, t*) and *CBFI*(*j, t*) are the cerebral blood flow index at channel *i* and *j* respectively, 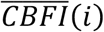 and 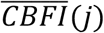 are the average CBFI over time, and *T* is the total number of datapoints in a time trace. Note that the number of possible combinations to calculate the correlation factor is 15, since we correlate two channels together out of six channels (6 choose 2).

**Fig 5.**
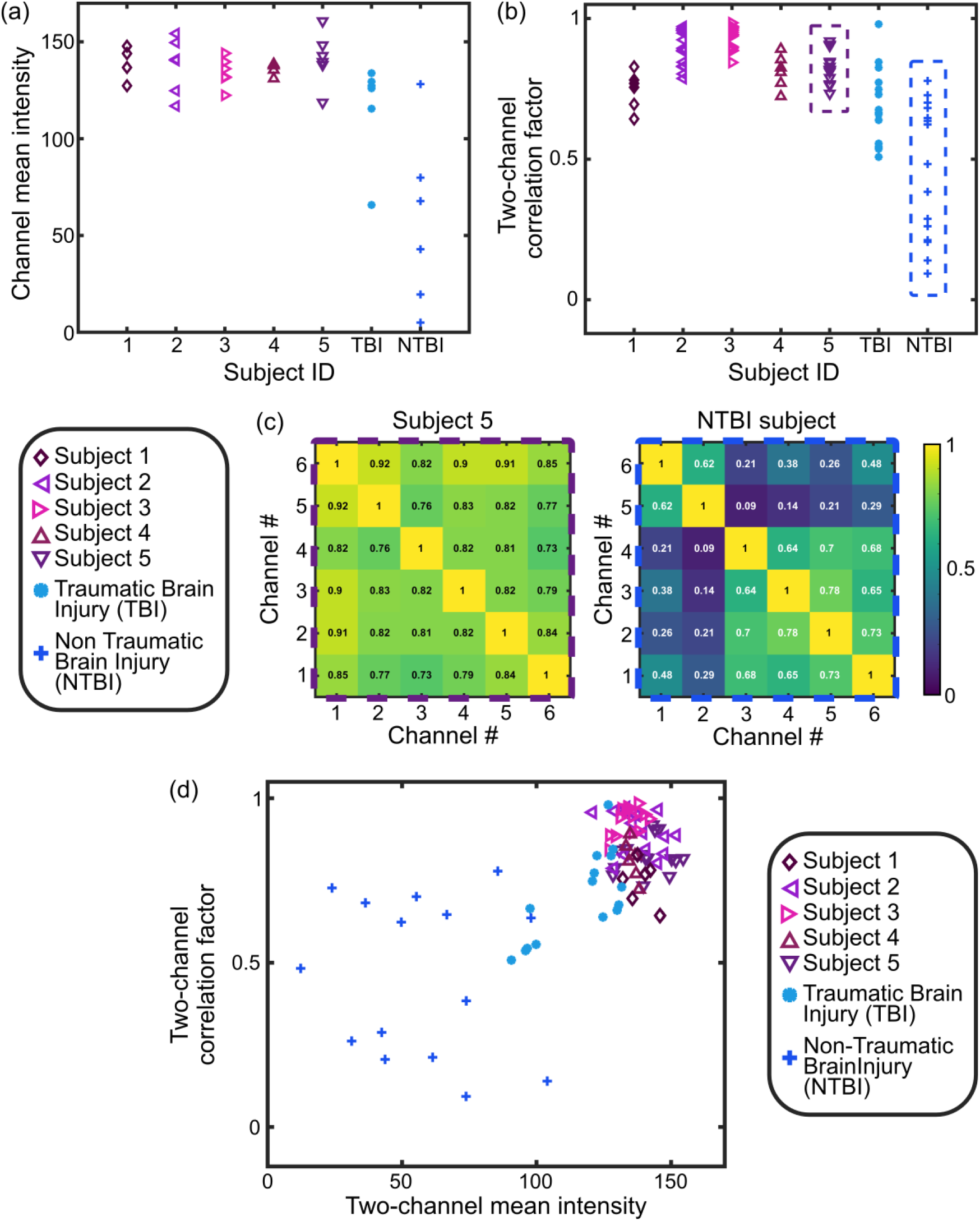
(a) Channel mean intensity and (b) two-channel correlation factor for each of the seven subjects. (c) Correlation matrix of all channels of Subject 5 (also shown in Fig. 4(a)) and NTBI Subject (also shown in Fig. 4(c)). (d) Two-channel correlation factor as a function of the two-channel mean intensity for all the subjects.

As shown in Figs. 5(a) and 5(b), both the channel mean intensity and two-channel correlation factor were significantly high for all the five healthy subjects. The skull implant subject exhibited similar channel mean intensity except for one channel but exhibited a lower correlation factor between channels. The NTBI subject however exhibited significantly lower mean intensity and correlation factor, especially on the four channels located on the back-right side of the head, i.e. the channels closer to the injury location.

Figure 5(c) presents the correlation matrix between all channels of Subject 5, corresponding to the CBF time traces shown in Fig. 4(a), and the NTBI subject, as shown in Fig. 4(c). While the correlation coefficients between all channels of Subject 5 are high (i.e., >0.75), the correlation coefficients between channels of the TBI subject reveal two distinct groups. Channels 1 and 2 are correlated with each other, while Channels 3 to 6 form a separate correlated group. This suggests that the channels positioned over the injury site exhibit distinct CBF time traces that are different compared to the other channels.

Figure 5(d) shows the two-channel correlation factor (Eq. (6)) as a function of the two-channel mean intensity. The two-channel mean intensity was calculated by averaging the mean intensity of the two channels used to compute the correlation factor. As shown, all subjects except the NTBI subject exhibited high correlation factors and mean intensities, indicating synchronized CBF and CBV dynamics with the same pulse waveform across all six channels for the subjects. In contrast, the NTBI subject displayed more variability in correlation factors and mean intensities, suggesting regional differences in CBF and CBV, likely due to the channels located on the injured region exhibiting different blood flow dynamics than the other channels.

It is important to note that these observations are preliminary due to the small sample size of only one of each TBI and NTBI subjects. A larger study with more brain injury subjects is necessary to draw conclusions about the effectiveness of our multi-channel approach for characterizing traumatic and non-traumatic brain injury. Nonetheless, this demonstrates the potential of the multi-channel SCOS utilizing its high throughput, multi-location, and parallel processing setup to compare signals among channels.

## 4 Conclusion

We demonstrated the effectiveness of the modular compact SCOS systems, expanded to six channels distributed around the head, for continuous monitoring of cerebral blood flow and blood volume at distinct regions of the brain over extended periods. The modular design of the system allows flexible positioning of the channels. As such, specific areas of interest can be easily accessed and monitored, with data processed in near real-time on a standard commercial laptop via a user-friendly GUI software. Battery-powered laser modules enhance the system’s portability, making it suitable for various environments, including field hospitals, ambulances, examination rooms or clinics. Aside from the camera cables, the entire system was integrated into a wearable headband that can be secured using zip ties or plastic Velcro straps. Optimized heat dissipation and insulation enable prolonged operation of the system for 30 minutes or longer while maintaining safe conditions of use and descent comfort level.

By comparing the six-channel CBF and CBV time traces across seven subjects with five healthy subjects, one subject who had mostly recovered from a TBI, and another with a severe non-traumatic brain injury (NTBI) with persistent symptoms, we observed noticeable differences in CBF and CBV dynamics at the site of worse structural damage compared to other locations. These findings were corroborated by MRI scans, where the locations of the channels showing altered CBF and CBV dynamics matched with the injury site in the MRI scans. The recovered TBI subject exhibited far less discrepancies between channels than the NTBI subject with persistent symptoms, and MRI scans showed relatively minor brain injury. These results underscore the potential of our multi-channel compact SCOS system to provide a rapid, non-invasive, and cost-effective preliminary diagnosis of brain injury. It is important to note that these findings are preliminary due to the small sample size. A larger study with more brain injury subjects is necessary to draw definitive conclusions about the effectiveness of our multi-channel approach for detecting the physiological sequelae of brain injury. As future research, we aim to extend the use of the system to a broader patient population, including individuals with TBI or non-traumatic (eg. cerebrovascular) events. By gathering more extensive and quantitative data, we hope to further validate the system’s diagnostic capabilities, providing physicians with a valuable and accessible tool for rapid preliminary diagnoses and treatment decisions.

## Data Availability

All data produced in the present study are available upon reasonable request to the authors

## Appendix A

Figure 2(a) of the main text showed the laser output power for three different power sources: a 9V DC bench power supply, a 9V alkaline battery, and a rechargeable 9V lithium-ion battery. We also tested zinc chloride 9V batteries, but the lasing lasted less than 5 minutes, making them unsuitable for use in the system. As shown, all three power sources provided the laser with an optical power of approximately 66 mW over time. The DC power supply maintained a constant output, while the alkaline and rechargeable lithium-ion batteries’ power decreased to zero after a few hours due to discharge.

We repeated those measurements over five different realizations and measured the duration of lasing. On average, the alkaline battery lasted an average of 3.2 ± 0.2 hours, and the rechargeable lithium-ion battery lasted 3.7 ± 0.2 hours. Larger instabilities occurred during the first 5 to 10 minutes where the laser power increased by about 3 mW due to the printed circuit board’s onset time (specifically, the charging/discharging of capacitors), the laser diode’s lasing dynamics, and the initial quick discharge of the battery’s voltage. After the onset period, the laser’s optical power remained stable at around 66 mW for all three power sources, with fluctuations of less than 0.6 mW for the alkaline battery, 0.4 mW for the rechargeable lithium-ion battery, and 0.3 mW for the DC power supply. We estimated that a laser power fluctuation of 0.5 mW or less during a recording is needed for stable measurements of CBF and CBV. Therefore, the system should be warmed up for 5 to 10 minutes before recording begins. To facilitate this, we have added a switch plug in front of the laser diode mount to allow the system to warm up while not exposing the subject with laser light.

We also measured the voltage of the power supply with an oscilloscope, Fig. 2(b), and observed that the battery discharge curves in Fig. 2(b) closely match the optical power curves in Fig. 2(a). The DC power curve remained constant without fluctuation, even in the first few minutes. Compared to the rechargeable lithium-ion battery, the alkaline battery had a larger discharge over the first 3.5 hours, explaining the higher stability of optical power with the rechargeable battery. The rechargeable lithium-ion batteries weighed on average 25 g, about 50% lighter than the alkaline batteries, which weighed on average 48 g. Due to their superior lasing stability, longer lasing period, and lighter weight, rechargeable lithium-ion batteries are more suitable for our compact SCOS system.

## Appendix B

To simplify the experiment and system implementation, measurements were conducted on hairless areas or regions with minimal hair, no more than a few millimeters long. The optical transmission of our system is optimal when both the light source and detector are positioned on areas with little to no hair, ideally, a hair-free square area of 1.5 cm^2^. While these hair-free areas are easily attainable for subjects with short hair, they are not for participants with longer hair. To address long hair interfering with system use, we plan on designing 3D-printed laser and camera mounts equipped with hair separators in the area of interest. This addition would minimize hair interference with the optical transmission process, and would enable the system to potentially be used on participants with longer hair.

## Conflict of Interest statement and Disclosures

The authors declare no conflicts of interest to declare. The human research protocol for this study received approval from the Caltech Committee for the Protection of Human Subjects and the Institutional Review Board (IRB), Caltech IRB Protocol IR21-1074.

## Acknowledgments

The authors thank Maya Dickson for her assistance during the design of the SCOS system and Professor Daniel Wagenaar for his help in designing the heat management system. This research was supported by the National Institutes of Health — Award No. 5R21EY033086-02 and USC Neurorestoration Center.

## Notes

### Competing Interest Statement

The authors have declared no competing interest.

### Author Declarations

Ethics committee/IRB of California Institute of Technology gave ethical approval for this work. The human research protocol for this study received approval from the Caltech Committee for the Protection of Human Subjects and the Institutional Review Board (IRB), Caltech IRB Protocol IR21-1074.

## References

1. P. McCrory et al., “Consensus statement on concussion in sport: the 4th International Conference on Concussion in Sport held in Zurich, November 2012,” Br J Sports Med 47(5), 250–258 (2013) [doi:10.1136/bjsports-2013-092313].

2. M. J. Ellis et al., “Neuroimaging Assessment of Cerebrovascular Reactivity in Concussion: Current Concepts, Methodological Considerations, and Review of the Literature,” Front. Neurol. 7 (2016) [doi:10.3389/fneur.2016.00061].

3. W. Schmid et al., “Review of wearable technologies and machine learning methodologies for systematic detection of mild traumatic brain injuries,” J. Neural Eng. 18(4), 041006 (2021) [doi:10.1088/1741-2552/ac1982].

4. C. C. Giza and D. A. Hovda, “The New Neurometabolic Cascade of Concussion,” Neurosurgery 75(Supplement 4), S24–S33 (2014) [doi:10.1227/NEU.0000000000000505].

5. C. C. Giza and D. A. Hovda, “The Neurometabolic Cascade of Concussion,” J Athl Train 36(3), 228– 235 (2001).

6. H. Phipps et al., “Characteristics and Impact of U.S. Military Blast-Related Mild Traumatic Brain Injury: A Systematic Review,” Front. Neurol. 11, 559318 (2020) [doi:10.3389/fneur.2020.559318].

7. E. Jones, N. T. Fear, and S. Wessely, “Shell Shock and Mild Traumatic Brain Injury: A Historical Review,” AJP 164(11), 1641–1645 (2007) [doi:10.1176/appi.ajp.2007.07071180].

8. J. F. Brundage et al., “Whither the ‘signature wounds of the war’ after the war: estimates of incidence rates and proportions of TBI and PTSD diagnoses attributable to background risk, enhanced ascertainment, and active war zone service, active component, U.S. Armed Forces, 2003-2014,” MSMR 22(2), 2–11 (2015).

9. A. D. Schweitzer et al., “Traumatic Brain Injury: Imaging Patterns and Complications,” RadioGraphics 39(6), 1571–1595 (2019) [doi:10.1148/rg.2019190076].

10. M. E. Haveman et al., “Predicting outcome in patients with moderate to severe traumatic brain injury using electroencephalography,” Crit Care 23(1), 401 (2019) [doi:10.1186/s13054-019-2656-6].

11. D. B. Hier et al., “Blood biomarkers for mild traumatic brain injury: a selective review of unresolved issues,” Biomark Res 9(1), 70 (2021) [doi:10.1186/s40364-021-00325-5].

12. K. Kawata et al., “Blood biomarkers for brain injury: What are we measuring?,” Neuroscience & Biobehavioral Reviews 68, 460–473 (2016) [doi:10.1016/j.neubiorev.2016.05.009].

13. S. D. Hicks et al., “Diagnosing mild traumatic brain injury using saliva RNA compared to cognitive and balance testing,” Clin Transl Med 10(6), e197 (2020) [doi:10.1002/ctm2.197].

14. A. I. R. Maas et al., “Traumatic brain injury: progress and challenges in prevention, clinical care, and research,” The Lancet Neurology 21(11), 1004–1060 (2022) [doi:10.1016/S1474-4422(22)00309-X].

15. S. Mahler et al., “Assessing depth sensitivity in laser interferometry speckle visibility spectroscopy (iSVS) through source-to-detector distance variation and cerebral blood flow monitoring in humans and rabbits,” Biomed. Opt. Express 14(9), 4964 (2023) [doi:10.1364/BOE.498815].

16. Y. X. Huang et al., “Compact and cost-effective laser-powered speckle contrast optical spectroscopy fiber-free device for measuring cerebral blood flow,” J. Biomed. Opt. 29(06), 067001 (2024) [doi:10.1117/1.JBO.29.6.067001].

17. C. G. Favilla et al., “Validation of the Openwater wearable optical system: cerebral hemodynamic monitoring during a breath-hold maneuver,” Neurophoton. 11(01) (2024) [doi:10.1117/1.NPh.11.1.015008].

18. B. Kim et al., “Measuring human cerebral blood flow and brain function with fiber-based speckle contrast optical spectroscopy system,” Commun Biol 6(1), 844 (2023) [doi:10.1038/s42003-023-05211-4].

19. T. Dragojević et al., “Compact, multi-exposure speckle contrast optical spectroscopy (SCOS) device for measuring deep tissue blood flow,” Biomed. Opt. Express 9(1), 322 (2018) [doi:10.1364/BOE.9.000322].

20. C. P. Valdes et al., “Speckle contrast optical spectroscopy, a non-invasive, diffuse optical method for measuring microvascular blood flow in tissue,” Biomed. Opt. Express, BOE 5(8), 2769–2784, Optica Publishing Group (2014) [doi:10.1364/BOE.5.002769].

21. Y. X. Huang et al., “Correlating stroke risk with non-invasive cerebrovascular perfusion dynamics using a portable speckle contrast optical spectroscopy laser device,” Biomedical Optics Express 15(10), 6083–6097 (2024) [doi:10.1364/BOE.534796].

22. Y. X. Huang et al., “Interferometric speckle visibility spectroscopy (iSVS) for measuring decorrelation time and dynamics of moving samples with enhanced signal-to-noise ratio and relaxed reference requirements,” Opt. Express 31(19), 31253 (2023) [doi:10.1364/OE.499473].

23. J. Xu et al., “Interferometric speckle visibility spectroscopy (ISVS) for human cerebral blood flow monitoring,” APL Photonics 5(12), 126102 (2020) [doi:10.1063/5.0021988].

24. J. Xu, A. K. Jahromi, and C. Yang, “Diffusing wave spectroscopy: A unified treatment on temporal sampling and speckle ensemble methods,” APL Photonics 6(1), 016105 (2021) [doi:10.1063/5.0034576].

25. Z. Dong et al., “Non-invasive laser speckle contrast imaging (LSCI) of extra-embryonic blood vessels in intact avian eggs at early developmental stages,” Biomed. Opt. Express 15(8), 4605 (2024) [doi:10.1364/BOE.530366].

26. G. E. Strangman, Z. Li, and Q. Zhang, “Depth Sensitivity and Source-Detector Separations for Near Infrared Spectroscopy Based on the Colin27 Brain Template,” PLoS ONE 8(8), e66319 (2013) [doi:10.1371/journal.pone.0066319].

27. F. B. Haeussinger et al., “Simulation of Near-Infrared Light Absorption Considering Individual Head and Prefrontal Cortex Anatomy: Implications for Optical Neuroimaging,” PLoS ONE 6(10), K. Hashimoto, Ed., e26377 (2011) [doi:10.1371/journal.pone.0026377].

28. C. Mansouri et al., “Depth sensitivity analysis of functional near-infrared spectroscopy measurement using three-dimensional Monte Carlo modelling-based magnetic resonance imaging,” Lasers Med Sci 25(3), 431–438 (2010) [doi:10.1007/s10103-010-0754-4].

29. S. A. Carp, M. B. Robinson, and M. A. Franceschini, “Diffuse correlation spectroscopy: current status and future outlook,” Neurophoton. 10(01) (2023) [doi:10.1117/1.NPh.10.1.013509].

30. W. Zhou et al., “Functional interferometric diffusing wave spectroscopy of the human brain,” Sci. Adv. 7(20), eabe0150 (2021) [doi:10.1126/sciadv.abe0150].

31. Laser Institute of America, American National Standard for Safe Use of Lasers, ANSI Z136.1-2014 (2014).

32. S. Zilpelwar et al., “Model of dynamic speckle evolution for evaluating laser speckle contrast measurements of tissue dynamics,” Biomed. Opt. Express 13(12), 6533 (2022) [doi:10.1364/BOE.472263].

33. M. B. Robinson et al., “Comparing the performance potential of speckle contrast optical spectroscopy and diffuse correlation spectroscopy for cerebral blood flow monitoring using Monte Carlo simulations in realistic head geometries,” Neurophoton. 11(01) (2024) [doi:10.1117/1.NPh.11.1.015004].

34. W. F. Sheppard, “On the Calculation of the most Probable Values of Frequency-Constants, for Data arranged according to Equidistant Division of a Scale,” Proceedings of the London Mathematical Society s1-29(1), 353–380 (1897) [doi:10.1112/plms/s1-29.1.353].

35. L. Kobayashi Frisk et al., “Comprehensive workflow and its validation for simulating diffuse speckle statistics for optical blood flow measurements,” Biomed. Opt. Express 15(2), 875 (2024) [doi:10.1364/BOE.502421].

36. C16 Committee, “Guide for Heated System Surface Conditions that Produce Contact Burn Injuries,” ASTM International [doi:10.1520/C1055-20].

37. International Electrotechnical Commission, “IEC 60601-1: Medical electrical equipment—Part 1: General requirements for basic safety and essential performance.,” International Electrotechnical Commission (2005).

38. R. Defrin et al., “Sensory determinants of thermal pain,” Brain 125(3), 501–510 (2002) [doi:10.1093/brain/awf055].

39. S. Yoo et al., “Responsive materials and mechanisms as thermal safety systems for skin-interfaced electronic devices,” Nat Commun 14(1), 1024 (2023) [doi:10.1038/s41467-023-36690-y].

40. H. Jeong et al., “Differential cardiopulmonary monitoring system for artifact-canceled physiological tracking of athletes, workers, and COVID-19 patients,” Sci. Adv. 7(20), eabg3092 (2021) [doi:10.1126/sciadv.abg3092].

41. J. Liu et al., “Enhancing wearable electronics through thermal management innovations,” Wearable Electronics 1, 160–179 (2024) [doi:10.1016/j.wees.2024.07.005].

